# Interest of seroprevalence surveys for the epidemiological surveillance of the SARS-CoV-2 pandemic in African populations: insights from the ARIACOV Project in Benin

**DOI:** 10.1101/2022.04.26.22274330

**Authors:** Parfait Houngbégnon, Odilon Nouatin, Anges Yadouleton, Benjamin Hounkpatin, Nadine Fievet, Eloïc Atindegla, Sebastien Deschavanne, Emilande Guichet, Ahidjo Ayouba, Raphaël Pelloquin, David Maman, Guillaume Thaurignac, Martine Peeters, Annonciat Aviansou, Salifou Sourakafou, Eric Delaporte, Achille Massougbodji, Gilles Cottrell

## Abstract

**Background:** Many SARS-CoV-2 seroprevalence surveys since the end of 2020 have disqualified the first misconception that Africa had been spared by the pandemic. Through the analysis of three SARS-CoV-2 seroprevalence surveys carried out in Benin as part of the ARIACOV project, we argue that the integration of epidemiological serosurveillance of the SARS-COV2 infection in the national surveillance package would be of great use to refine the understanding of the COVID-19 pandemic in Africa.

**Methods:** Three repeated cross-sectional surveys have been carried out in Benin, two in Cotonou, the economic capital in March and May 2021, and one in Natitingou, a semi-rural city in North in August 2021. The global and by age-groups weighted seroprevalences have been estimated and the risk factors of the infection by SARS-COV-2 have been assessed by using logistic regression.

**Results:** In Cotonou, a slight increase in overall age-standardized SARS-CoV-2 seroprevalence from 29.77% (95% CI: 23.12-37.41%) at the first survey to 34.86% (95% CI: 31.57-38.30%) at the second survey was observed. In Natitingou the global adjusted seroprevalence was 33.34% (95% CI: 27.75-39.44%), much higher than expected. Adults over 40 seemed to be more at risk than the youngest during the first survey in Cotonou but no longer in the second survey, showing the persistence of the SARS-COV-2 virus circulation outside the epidemic waves.

**Conclusions:** A routine serological surveillance on strategic sentinel sites and / or populations could constitute a cost / effective compromise to better anticipate the onset of new waves and define public health strategies.

## Introduction

In Benin, the first case of SARS-CoV-2 infection was detected March, 16th 2020 (1). From the very first months of the COVID-19 pandemic, most of African countries implemented public health measures (2,3) to counter the spread of the virus, in particular sanitary cords, border closures and sometimes containment. These health governmental decisions were accompanied by sensitization campaigns on the wearing of a mask, social distancing measures, barrier gestures, hand washing, and also by increasing the capacities of national laboratories in terms of SARS-CoV-2 infection screening by Polymerase Chain Reaction (PCR) (4). The objective of this strategy was to interrupt the chains of transmission as quickly as possible, knowing that the capacity to handle severe cases in hospital was limited in most countries. Nevertheless, the number of tests carried out in Africa has remained low compared to the rest of the world (5,6) and most often mainly reserved for capitals or large cities, making it difficult to assess the reality of the circulation of the virus in country sides. So far, the severity of the disease has been less than in other parts of the world (7–9) and the proportion of asymptomatic or pauci-symptomatic cases among infected people is probably very high. As a result, data on the number of COVID-19 cases in Africa are scarce (10,11) and their ability to reflect the reality of the extent of the pandemic is questionable. The low number of detected cases and reported deaths in Africa (12–19) has sometimes been interpreted as the fact that the continent had been spared from the pandemic. However, many SARS-CoV-2 seroprevalence surveys (following the WHO recommendations (20)) published since the end of 2020 in different African countries, have now definitively disqualified this misconception (21). The ARIACOV project (Support for the African Response against COVID-19) which carried out SARS-CoV-2 seroprevalence surveys in 6 West and Central African countries (Benin, Ghana, Cameroon, Democratic Republic of Congo, Guinea and Senegal) together with other initiatives helped to demonstrate that the virus has circulated widely on the continent (4,11,21,22). All of these results show that, as highly useful as it is to understand the general dynamics of the COVID-19 pandemic, routine epidemiological surveillance in Africa exclusively based on the detection of infected cases by PCR (or antigenic RDT more recently) has proved unable to account for the true extent of the epidemic in the African context (limited screening capacity, proportion of asymptomatic cases probably overwhelming). The integration of epidemiological sero-surveillance based on the detection of antibodies against SARS-CoV-2 in the national surveillance package would certainly be of great use to refine the understanding of the dynamics of the epidemic and to detect the most at risk of infection groups in the populations. The aim of this work is to argue this idea through the analysis of three SARS-CoV-2 seroprevalence surveys carried out in Benin as part of the ARIACOV project.

## Methods

### Study area

The first two surveys of the study were conducted in Cotonou, the economic capital of Benin with the port and the airport, with an area of 79 km2 and an estimated population of 840 313 in 2020 (23). To assess the COVID-19 epidemic situation in a city where epidemic activity was assumed to be low according to the national surveillance, the third survey was carried out at Natitingou, a middle-size city located more than 500 kilometers at North of Benin. Natitingou has an area of 3045 km2 with an estimated population of 128 511 in 2020 (23).

### Study design and participants

We conducted three cross-sectional SARS-CoV-2 seroprevalence studies in households. The first and the second studies took place in Cotonou, in March 3 to 15, 2021 (decrease of the second wave), and May 27 to June 8, 2021(between the second and the first wave), respectively. The third study took place in Natitingou from August 20 to 26, 2021 (increase of the third wave). These studies were carried out as part of the ARIACOV project and used the WHO population-based age stratified seroepidemiological investigation protocol for COVID-19 infection version 2.0.

A two-stage cluster sampling was employed in each study of Cotonou. At the first stage, 50 neighborhoods among 143 were randomly drawn with a probability proportional to their population size, and at the second stage 12 households were selected per neighborhood. In order to balance the age groups (to be able to compare the 2 surveys by age groups), in half of the households all the residents were invited to participate in the study and in the remaining 50%, only the residents aged 40 years and older were invited to participate. In Natitingou, a two-stage cluster sampling was also carried out, where 30 neighborhoods among 77 were randomly drawn with a probability proportional to their size, and then 5 households were randomly drawn in each neighborhood. In Natitingou, all the residents of any household were invited to participate in the study. In each study, the eligible participants were proposed to fill an electronic questionnaire aiming to collect on smartphones general (age, gender…) and socio-enonomic (occupation, individual family home, common yard…) data, as well as experience of COVID-19 confirmed disease or related symptoms in the past 3 months and/or hospitalization (defined asymptomatic if no symptom). The participants were eventually proposed to be performed a blood sample for SARS-CoV-2 serological analysis in the laboratory.

### Ethical considerations

Ethics approval (N°131/MS/DRFMT/CNERS/SA) and Statistical approval (N°26/2020/MPD/INSAE/DCSFR) were obtained respectively from the Comité d’Ethique pour la Recherche en Santé and the Conseil National de la Statistique. All residents (adults and children) were informed about the study objectives and procedures. Adults provided written consent to participate in the study and to be tested for SARS-CoV-2 serology prior to starting the interview. Written parental consent and children assent were obtained prior to enrollment of young participants. At the end of the study, each participant was informed about his serology status.

### Antibody detection to SARS-CoV-2/COVID-19

Venous blood samples (2-5ml) collected in EDTA tube from participants were carefully transported to the laboratory of the Clinical Research Institute of Benin (IRCB). After centrifugation and inactivation at 56°C during 45 min, plasmas samples were used for SARS-CoV-2 serological analyses. In this study, the Luminex-based assay was used to simultaneously detect IgG antibodies to two viral antigen, i.e recombinant nucleocapsid (NC) and spike (SP) proteins of SARS-CoV-2, as previously described (24). This dual target strategy for SARS-CoV-2 IgG antibodies detection give a great sensitivity (100%) and specificity (99,7%) that was previously validated on an panel of more than 1 000 samples from Africa (including 160 samples from Benin) before COVID-19 (22,24). Results were expressed as the median fluorescence intensity (MFI) for 100 beads. Cutoff values were also previously determined using pre-COVID-19 samples and those from COVID-19 positive and hospitalized patients (24). An experiment was validated when (1) the MIF value of the blank is less than 50 MFI regardless of the antigen and (2) the MFI value of the negative control is less than 100. A sample is considered positive for anti-SARS-CoV-2 IgG when positive both for anti-SP IgG and anti-NP IgG, and is declared negative when negative for this both antigens.

### Statistical analysis

The statistical analysis was performed using STATA 14 (StataCorp, College Station, TX, USA). In Cotonou the weighted estimations of the SARS-CoV-2 seroprevalence (and 95% confidence intervals) were performed in the 2 surveys by age-standardizing the crude seroprevalence using available national census data (23). We estimated the overall SARS-CoV-2 seroprevalence and the seroprevalence stratified by sex and age groups in each of the 3 surveys. A weighted multivariable logistical regression was performed to determine the factors associated with positive serology anti-SARS-COV-2. The age group and symptom variables were forced into the model.

## Results

### Study population

In Cotonou, during the first and second surveys, 617 and 606 households were visited respectively, with a total of 1596 and 1842 eligible individuals respectively (Table 1). Among them, 1416 (89%) and 1465 (80%) individuals were present and agreed to participate in the first and second survey respectively and finally, 1405 and 1460 compliant blood samples with epidemiological data were collected, respectively. The average size of the households surveyed was 3.26 ± 1.98 for the first survey and 3.97 ± 1.81 for the second survey. In Natitingou, 155 households were visited for a total of 815 eligible individuals, 742 (91%) individuals were present and agreed to participate in the study, of which 735 did provide a compliant blood sample. The average size of the households surveyed was 6.06 ± 2.16. The general characteristics of the households and participants are described in Table 1.

**Tableau 1 :**
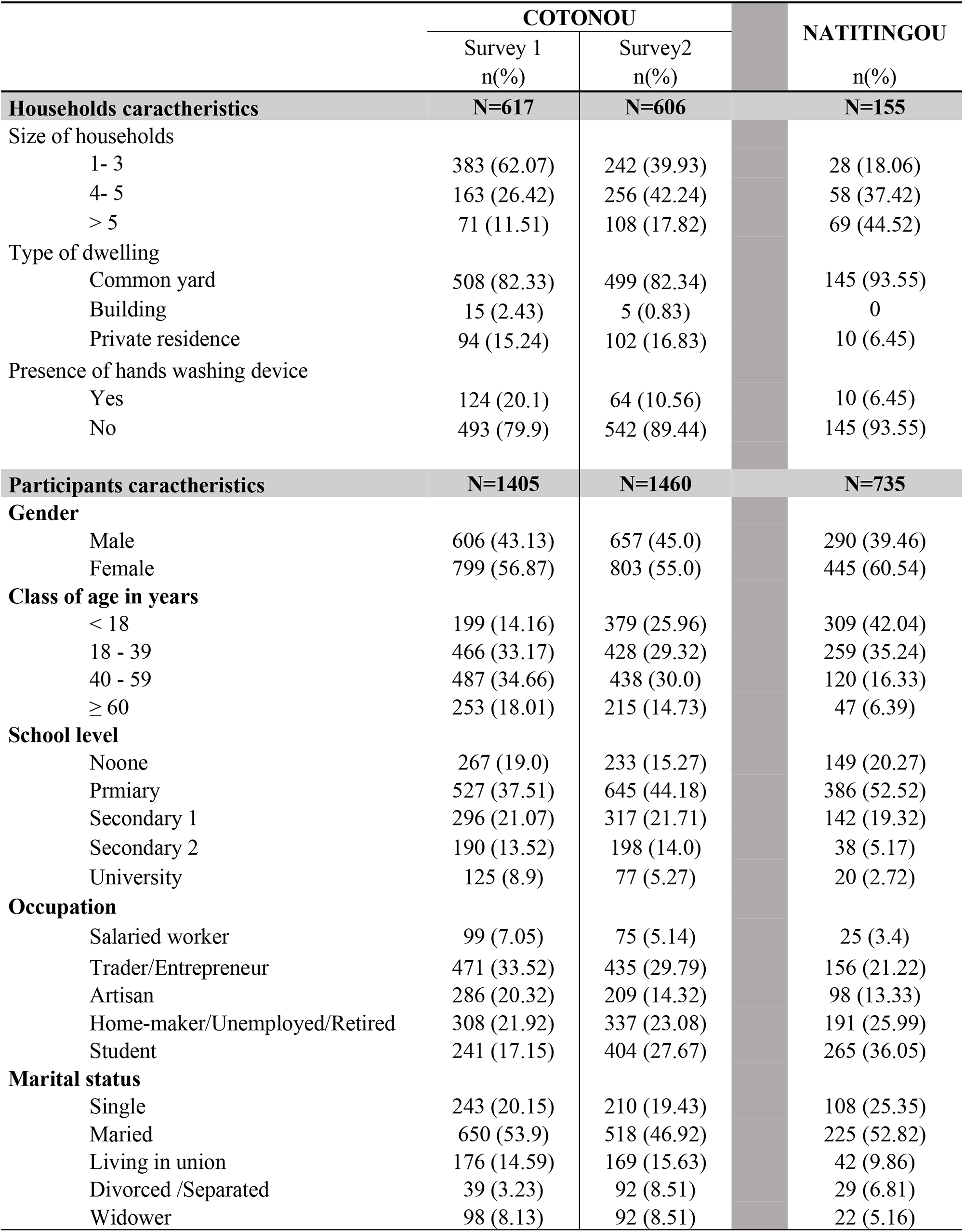
Characteristics of households and participants surveyed in Cotonou and Natitingou

|  | COTONOU |  | NATITINGOU |
| --- | --- | --- | --- |
|  | Survey 1<br>n(%) | Survey2<br>n(%) | n(%) |
| **Households characteristics** | **N=617** | **N=606** | **N=155** |
| Size of households |  |  |  |
| 1- 3 | 383 (62.07) | 242 (39.93) | 28 (18.06) |
| 4- 5 | 163 (26.42) | 256 (42.24) | 58 (37.42) |
| > 5 | 71 (11.51) | 108 (17.82) | 69 (44.52) |
| Type of dwelling |  |  |  |
| Common yard | 508 (82.33) | 499 (82.34) | 145 (93.55) |
| Building | 15 (2.43) | 5 (0.83) | 0 |
| Private residence | 94 (15.24) | 102 (16.83) | 10 (6.45) |
| Presence of hands washing device |  |  |  |
| Yes | 124 (20.1) | 64 (10.56) | 10 (6.45) |
| No | 493 (79.9) | 542 (89.44) | 145 (93.55) |
| **Participants characteristics** | **N=1405** | **N=1460** | **N=735** |
| **Gender** |  |  |  |
| Male | 606 (43.13) | 657 (45.0) | 290 (39.46) |
| Female | 799 (56.87) | 803 (55.0) | 445 (60.54) |
| **Class of age in years** |  |  |  |
| < 18 | 199 (14.16) | 379 (25.96) | 309 (42.04) |
| 18 - 39 | 466 (33.17) | 428 (29.32) | 259 (35.24) |
| 40 - 59 | 487 (34.66) | 438 (30.0) | 120 (16.33) |
| ≥ 60 | 253 (18.01) | 215 (14.73) | 47 (6.39) |
| **School level** |  |  |  |
| Noone | 267 (19.0) | 233 (15.27) | 149 (20.27) |
| Prmiary | 527 (37.51) | 645 (44.18) | 386 (52.52) |
| Secondary 1 | 296 (21.07) | 317 (21.71) | 142 (19.32) |
| Secondary 2 | 190 (13.52) | 198 (14.0) | 38 (5.17) |
| University | 125 (8.9) | 77 (5.27) | 20 (2.72) |
| **Occupation** |  |  |  |
| Salaried worker | 99 (7.05) | 75 (5.14) | 25 (3.4) |
| Trader/Entrepreneur | 471 (33.52) | 435 (29.79) | 156 (21.22) |
| Artisan | 286 (20.32) | 209 (14.32) | 98 (13.33) |
| Home-maker/Unemployed/Retired | 308 (21.92) | 337 (23.08) | 191 (25.99) |
| Student | 241 (17.15) | 404 (27.67) | 265 (36.05) |
| **Marital status** |  |  |  |
| Single | 243 (20.15) | 210 (19.43) | 108 (25.35) |
| Maried | 650 (53.9) | 518 (46.92) | 225 (52.82) |
| Living in union | 176 (14.59) | 169 (15.63) | 42 (9.86) |
| Divorced /Separated | 39 (3.23) | 92 (8.51) | 29 (6.81) |
| Widower | 98 (8.13) | 92 (8.51) | 22 (5.16) |

### SARS-CoV-2 seroprevalence in Cotonou and Natitingou

In Cotonou, a slight increase in overall age-standardized SARS-CoV-2 seroprevalence from 29.77% (95% CI: 23.12-37.41%) at the first survey to 34.86% (95% CI: 31.57-38.30%) at the second survey was observed (Table 2). In Natitingou, the global adjusted seroprevalence was also as high as 33.34% (95% CI: 27.75-39.44%). The seroprevalence stratified by sex is presented Table 2.

**Table 2 :** Adjusted seroprevalence of SARS-CoV-2 in Cotonou and Natitingou

|  | COTONOU<br>Survey 1 | COTONOU<br>Survey 2 |  | NATITINGOU |
| --- | --- | --- | --- | --- |
| Adjusted seroprevalence in all the population<br>95% of CI | 29.77%<br>[23.12 – 37.41] | 34.86%<br>[31.57– 38.30] |  | 33.34%<br>[27.75 – 39.44] |
| Adjusted seroprevalence in the **women** population<br>95% of CI | 30.72%<br>[22.49 – 40.38] | 38.08%<br>[33.53 – 42.87] |  | 34.93%<br>[29.24 – 41.10] |
| Adjusted seroprevalence in the **men** population<br>95% of CI | 28.64%<br>[22.05 – 36.28] | 31.50%<br>[27.63 – 35.65] |  | 30.99%<br>[22.96 – 40.36] |

The proportion of symptomatic subjects according to the SARS-COV-2 serology is shown Figure 1. The first survey in Cotonou showed a lower seroprevalence among the youngest age groups (less than 40 years) compared to 40 years and above (Figure 2), but this trend was no longer observed at the second survey. In Natitingou, a higher seroprevalence was observed among people aged over 60 years or over but the sample size in this age group was quite low (Figure 3).

**Figure 1 :**
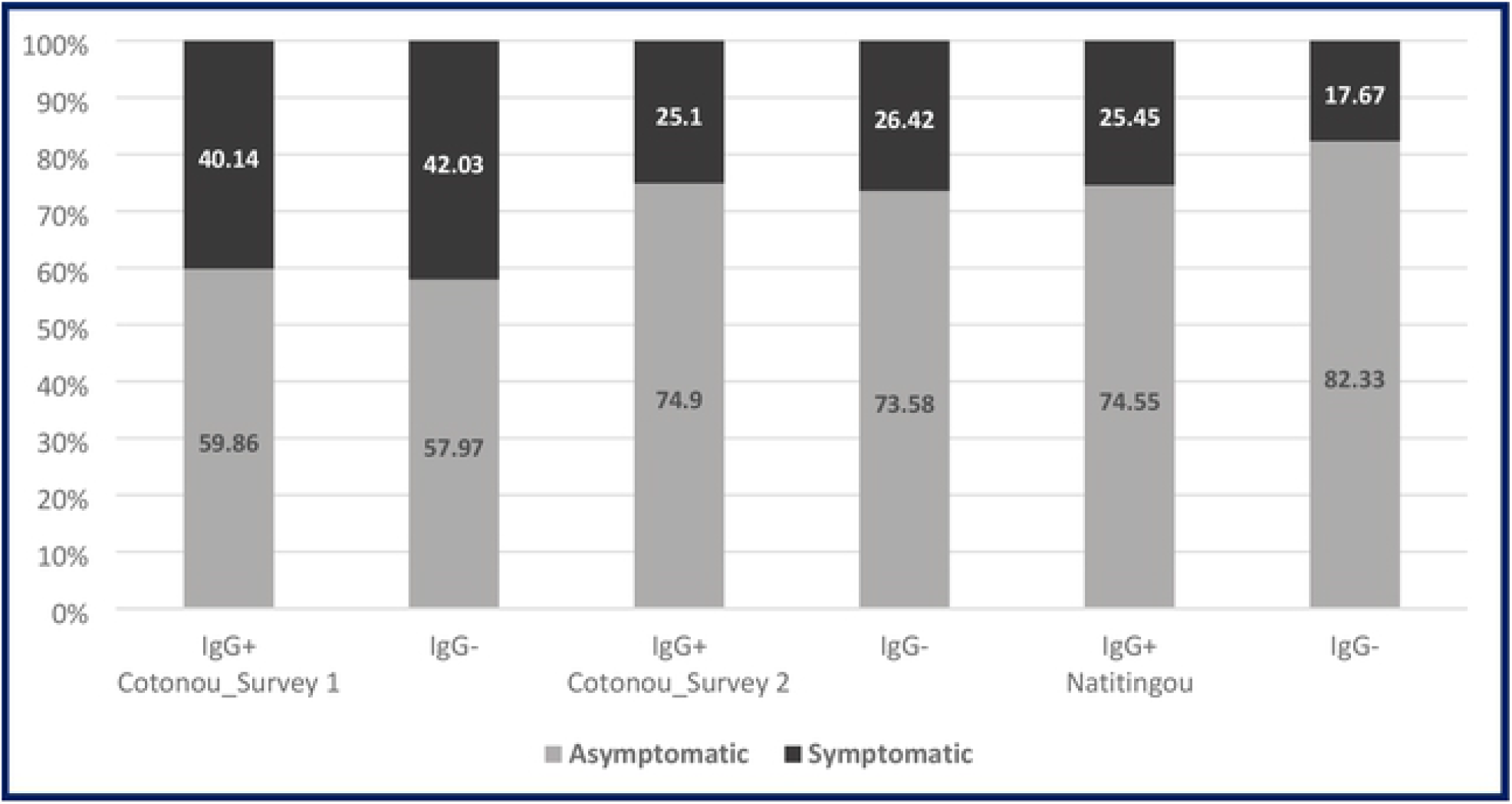
Proportion of Asymptomatic and Symptomatic by IgG results

**Figure 2 :**
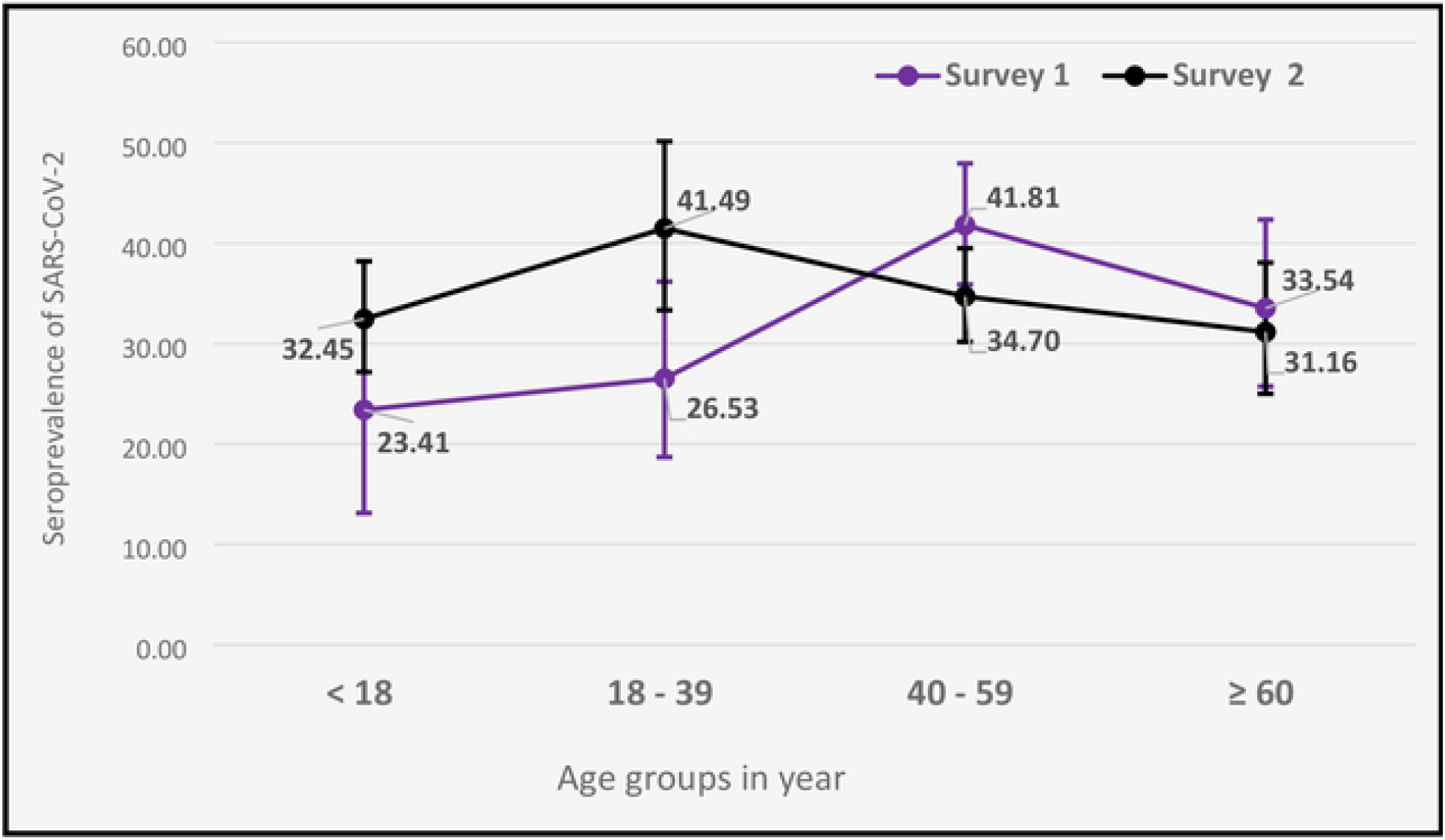
Seroprevalence of SARS-CoV-2 according to age groups in Cotonou in the first and second surveys (March and May 2021)

**Figure 3 :**
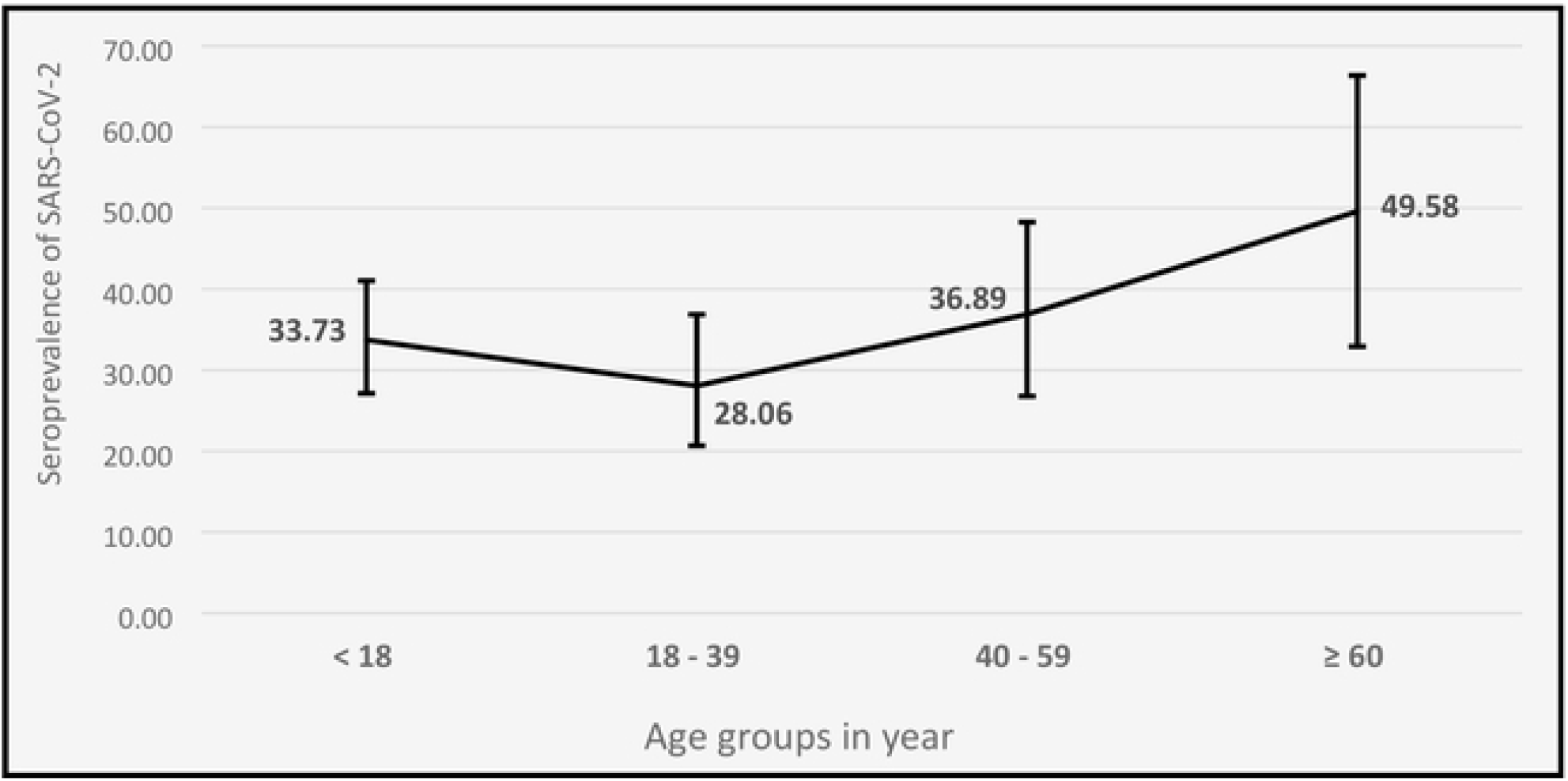
Seroprevalence of SARS-CoV-2 according to age groups in Natitingou (August 2021)

### SARS-CoV-2 risk factors in Cotonou and Natitingou

The multivariate logistic regression (Table 3) presents the covariates for which a clear association with a SARS-CoV-2 seropositivity was found in at least one of the surveys. The three surveys showed an OR greater than 1 in women (higher risk of positive serology, with a p <0.05 only for the second Cotonou survey). A consistent trend towards a higher risk in more populated households or in households living in common yards (vs. more isolated structures) was observed during the first survey in Cotonou, but this was not found in the others surveys. Adults over 40 seemed to be more at risk than the youngest (with p> 0.05 however) during the first survey in Cotonou, while in the second survey the ORs (and 95% CI) in the different age groups were more homogeneous. In Natitingou, a clear tendency for increased risk with age was found, with an OR of 2.67 for people aged 60 and over (p <0.05). No association was found between symptomatic status during the last 3 months and SARS-CoV-2 positive serology in surveys in Cotonou. In Natitingou, an incread-risk OR in symptomatic people was observed (OR = 1.33), although this association was not very clear (p = 0.25).

**Tableau 3 :**
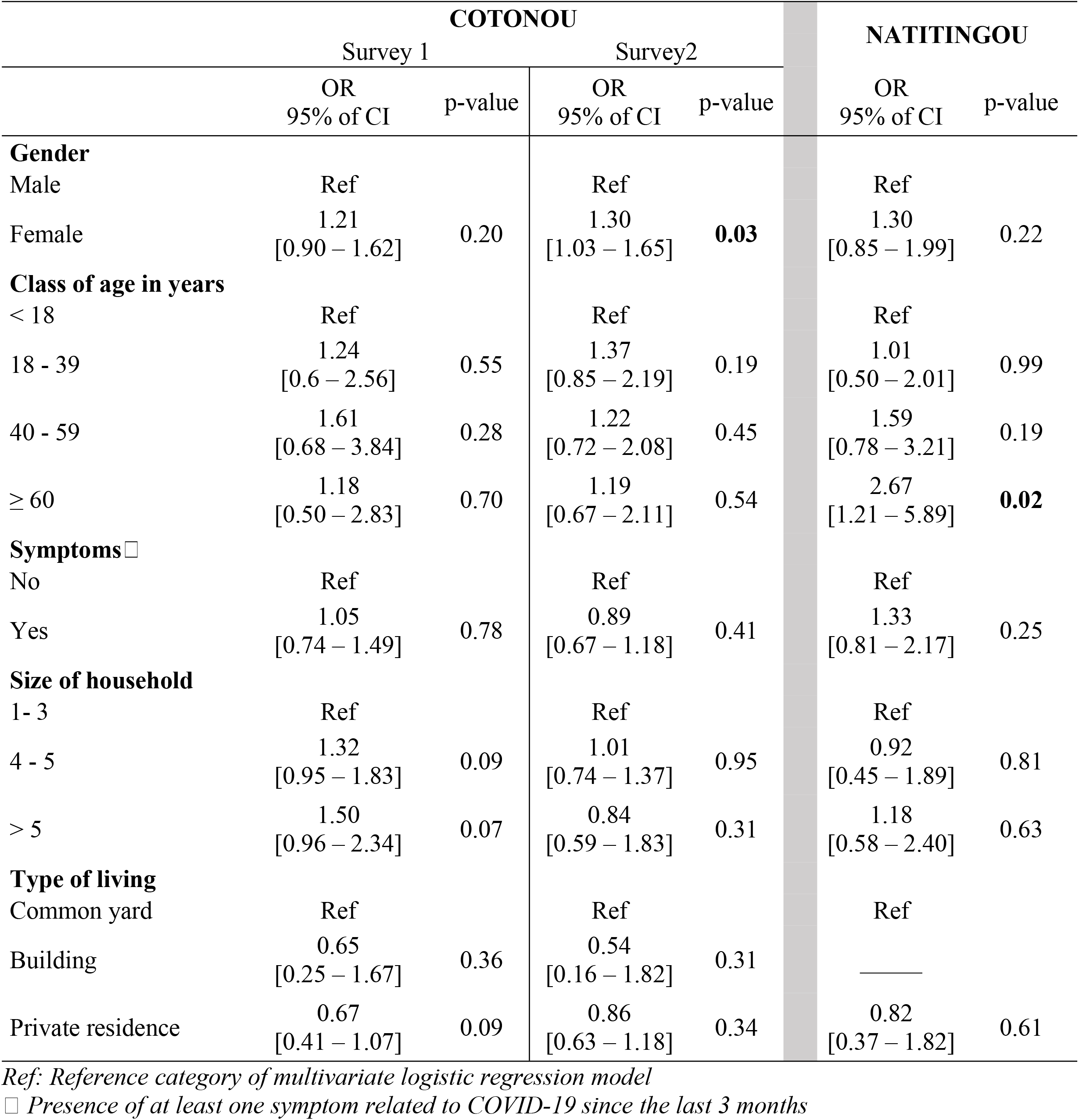
Multivariate risk factors analysis for SARS-CoV-2 seropositivity

## Discussion

We presented three cross-sectional seroprevalence studies in Benin, two carried out in Cotonou in March and May-June 2021, the third in Natitingou (a medium-sized city in the north of Benin) in August 2021. The main result was that the estimated prevalence in the 3 surveys turned out to be much higher than shown by virological surveillance based on PCR tests. For example, based on the available national demographic data (23), the estimated number of people having been in contact with the virus from the first survey in Cotonou amounts to 250 161 ([194 289; 314 361]) for an estimated population of around 840,000 inhabitants living in Cotonou whereas just after the first survey in Cotonou, the number of cases diagnosed by PCR nationwide was 6 501 (only one case detected for every 40 infections). This ratio is in accordance with numerous seroprevalence surveys carried out in Africa (4,11,25,26).The seroprevalences we found in our 3 surveys fit also well with a recent meta-analysis showing a combined seroprevalence of 25% (95% C.I. [13% - 39%] in West Africa (21).

Some studies, for example in Mali or in Zimbabwe have measured seroprevalences before and after epidemic waves, demonstrating the substantial impact of these waves on seroprevalence estimates (11,27). Unlike these studies, the two surveys in Cotonou were both carried out during and after second wave (end of January to end of March) and only 2 months apart. This largely explains the smaller increase in seroprevalence between the two surveys (from 29.77% to 34.86%). Interestingly in the second survey in Cotonou, the prevalence among the youngest caught up with the prevalence of the older age groups. This could suggest an impact of the sensitization discourse (particularly oriented towards older age groups), associated to a greater respect for distancing measures between the two surveys by older age groups compared to younger ones. It reflects also the persistence of virus circulation even outside of waves, so even when virological PCR monitoring does not seem to suggest any increase in COVID-19 cases incidence.

Based on the findings in Cotonou, our hypothesis was that the prevalence of infected people with the SARS-CoV-2 virus was undoubtedly underestimated throughout the country. The goal of the investigation in Natitingou was then to assess the situation in an area where available data suggested very low circulation of the virus, despite the start of the third COVID-19 epidemic wave in Benin (August to October 2021). The estimation of age-groups prevalences was not a main objective, which justifies the non age-group stratified design in Natitingou. Our hypothesis was confirmed, with an estimated overall adjusted prevalence of 33.34%, well above the low PCR positive rate detected by the epidemiological surveillance in the study area (data not shown), and in agreement with the high prevalence observed in semi-rural areas after the second wave in Mali for example (26).

Across the three surveys, the presence of symptoms in the past 3 months has not shown any relationship with positive serology, as observed in several other studies (4,28–31). This supports the hypothesis that the vast majority of the infected people have been asymptomatic or pauci-symptomatic, even if the information regarding the symptoms may be affected by a memorization bias. The proportion of people (even symptomatic) having tested for COVID-19 was very low (less than 5%, data not shown), which shows the limits of surveillance based on PCR tests intended for people presenting spontaneously.

Our study has limitations. In Cotonou, the design was repeated cross-sectional rather than longitudinal surveys. Consequently, the differences between the prevalence by age groups in the two surveys may be because the samples were carried out different neighborhoods of Cotonou. Nonetheless, the relatively small increase in prevalence between the two surveys is consistent with the relatively narrow time interval (2 months) with no epidemic wave in between. Another limitation is that in Natitingou the design does not allow to estimate the prevalences by age group as precisely as in Cotonou. However, all age groups seem to be highly affected, which is consistent with what was observed in Cotonou and in other studies including the ARIACOV project (11).

All of these results show the essential value of seroprevalence surveys in providing a more accurate view of the spread and the extent of the COVID19 epidemic than the reported cases from virological PCR-based surveillance only. Virological surveillance seems suitable for describing epidemic waves when they are in progress, but turns out to be inadequate neither for capturing a precise quantitative estimate of the extent and spread of the epidemic in populations. It fails also to apprehend clearly the persistence of the virus and its basal dynamics between epidemic waves. Our results show, along with others in Africa, that a more systematic use of serological surveillance to support an overall strategy of routine epidemiological surveillance would certainly be useful in overcoming these shortcomings. The interest of associating seroprevalence studies with PCR (or RDT) screening as part of the epidemiological surveillance of the pandemic was pointed out in early 2020 by international health institutions such as the World Health Organization (32) or the Center of Disease Control and Prevention (33) especially given the high proportion of infected people with few or no symptoms. Their rapid integration into the national packages for monitoring the pandemic in northern countries has effectively made it possible to better estimate the real extent of the transmission of the virus, the risk factors for infection and the foci of transmission. In particular before mass vaccination of populations, knowledge of the true proportion and sub-groups of the population with anti-SARS-COV-2 antibodies is an essential element to better guide the national vaccine strategies. These considerations are particularly important in an African context where many countries are constrained to lighten, sometimes significantly, their epidemiological surveillance systems because of its logistical burden and high cost, especially between epidemic waves when the epidemic situation seems stable. However, as shown here, the virus probably continues to diffuse quietly, contributing to the emergence of new variants and new waves, particularly in populations that are still poorly vaccinated. It is therefore urgent to think about a way to maintain a routine epidemiological surveillance adapted to this context. To minimize the cost, a routine serological surveillance on strategic sentinel sites and / or populations could constitute a cost / effective compromise to better anticipate the onset of new waves and characterize the most vulnerable subgroups of populations. In order to take into account the logistical constraints, the use of an easy-to-use diagnostic tool such as serological RDTs, well known to health workers as well as to populations, as it is already used for the COVID-19 case detection (antigenic RDTs) and in other pathologies like malaria, could prove interesting.

## Data Availability

According to the Benin law (the Authority of Personal Data Protection), we'll not be able to share the data of this publication.

## Acknowledgements

Special gratitude to all the study participants. We acknowledge all the technicians and the field surveyors. We highlighted the facilities offer by the administrative and health authorities at central and local levels. We acknowledge Inès Boko, Elisée ADIMI, Romuald AKOHO and Jean-François Etard for their appreciated contribution to the study.

